# Retinal BioAge Reveals Indicators of Cardiovascular-Kidney-Metabolic Syndrome in US and UK Populations

**DOI:** 10.1101/2024.07.18.24310670

**Authors:** Ehsan Vaghefi, Songyang An, Shima Moghadam, Song Yang, Li Xie, Mary K. Durbin, Huiyuan Hou, Robert N. Weinreb, David Squirrell, Michael V. McConnell

## Abstract

**Background:** There is a growing recognition of the divergence between biological and chronological age, as well as the interaction among cardiovascular, kidney, and metabolic (CKM) diseases, known as CKM syndrome, in shortening both lifespan and healthspan. Detecting indicators of CKM syndrome can prompt lifestyle and risk-factor management to prevent progression to adverse clinical events. In this study, we tested a novel deep-learning model, retinal BioAge, to determine whether it could identify individuals with a higher prevalence of CKM indicators compared to their peers of similar chronological age.

**Methods:** Retinal images and health records were analyzed from both the UK Biobank population health study and the US-based EyePACS 10K dataset of persons living with diabetes. 77,887 retinal images from 44,731 unique participants were used to train the retinal BioAge model. For validation, separate test sets of 10,976 images (5,476 individuals) from UK Biobank and 19,856 retinal images (9,786 individuals) from EyePACS 10K were analyzed. Retinal AgeGap (retinal BioAge – chronological age) was calculated for each participant, and those in the top and bottom retinal AgeGap quartiles were compared for prevalence of abnormal blood pressure, cholesterol, kidney function, and hemoglobin A1c.

**Results:** In UK Biobank, participants in the top retinal AgeGap quartile had significantly higher prevalence of hypertension compared to the bottom quartile (36.3% vs. 29.0%, p<0.001), while the prevalence was similar for elevated non-HDL cholesterol (77.9% vs. 78.4%, p=0.80), impaired kidney function (4.8% vs. 4.2%, p=0.60), and diabetes (3.1% vs. 2.2%, p=0.24). In contrast, EyePACS 10K individuals in the top retinal AgeGap quartile had higher prevalence of elevated non-HDL cholesterol (49.9% vs. 43.0%, p<0.001), impaired kidney function (36.7% vs. 23.1%, p<0.001), suboptimally controlled diabetes (76.5% vs. 60.0%, p<0.001), and diabetic retinopathy (52.9% vs. 8.0%, p<0.001), but not hypertension (53.8% vs. 55.4%, p=0.33).

**Conclusion:** A deep-learning retinal BioAge model identified individuals who had a higher prevalence of underlying indicators of CKM syndrome compared to their peers, particularly in a diverse US dataset of persons living with diabetes.

**Clinical Perspective:** *What Is New?:*

- Accelerated biological aging predicted by a novel deep-learning analysis of standard retinal images was able to detect multiple indicators of the new cardiovascular-kidney-metabolic syndrome in US and UK populations.

*What Are the Clinical Implications?:*

- Rapid, point-of-care analysis of images from routine eye exams can broaden access to the detection and awareness of adverse cardiovascular, kidney, and metabolic health.
- With the broad range of prevention interventions to reduce progression of cardiovascular-kidney-metabolic syndrome, earlier and broader detection is important to improve public health outcomes.

## Introduction

Cardiovascular, kidney, and metabolic diseases are major contributors to global morbidity and mortality, and healthcare costs.^1–4^ There is also a growing recognition of the pathophysiologic overlap of these conditions and their interplay in risk for adverse clinical events. These combined conditions have been described recently as the cardiovascular-kidney-metabolic (CKM) syndrome by the American Heart Association, with a staging system based on the presence and severity of indicators for all three diseases.^5–6^ Thus, detecting indicators of CKM syndrome presents an opportunity to enhance interventions for prevention and risk-factor management to attenuate progression to cardiovascular events, kidney failure, and diabetes complications.

The retina is unique in being the only part of the human vasculature that is directly visible, and it is widely recognized that the risk for, and severity of, important systemic diseases can be detected from retinal images.^7^ These include the well-known retinal signs of hypertensive and diabetic retinopathy.^8,9^ With progress in artificial intelligence (AI), and deep learning (DL) in particular, there are now many studies using AI to analyze retinal images.^10,11^ We and others have shown that these DL models can detect cardiovascular risk, chronic kidney disease, and diabetic retinopathy.^12–21^

DL models analyzing retinal images have also been developed to estimate “biological age.” Since its initial inception in the late 1960s, there is increasing recognition that biological age - the concept that individuals age at different rates - impacts both the risk of an individual developing a major disease and the severity to which they are impacted by it.^22–25^ That individuals age at different rates is not captured by traditional regression-based risk equations as they assume all individuals in the population age at the same rate. Thus, a retinal biological age may provide a broader indicator of health for an individual. We and others have shown previously that biological age derived from AI analysis of retinal images correlates with telomere length and higher cardiovascular and cancer mortality.^26–29^ Here we apply our retinal BioAge DL model to detect indicators of CKM syndrome in two different cohorts – a UK-based general population cohort and a diverse, US-based cohort of persons living with diabetes.

## Methods

### Retinal BioAge model development

The retinal BioAge DL model was developed as an extension of prior work determining atherosclerotic cardiovascular disease (ASCVD) risk from retinal images.^13^ The retinal BioAge was derived by comparing the AI-predicted 10-year ASCVD risk score of an individual to those of their peers of higher and lower chronological age, as published previously.^26^ Briefly:

- The degree of similarity of the individual’s AI-predicted ASCVD risk score with the ASCVD risk score obtained from other similar-aged people was determined.
- The average (chronological) age of that individuals’ closest neighbors, based on the condition of the retina as expressed by multi-dimensional features extracted from the DL models, was determined.
- The mean expected age for a person with the individual’s AI-predicted ASCVD risk was determined.
- These data were then combined in a probabilistic manner to give a final value for the retinal BioAge.
- The equation for calculating retinal BioAge is provided in the Supplementary Methods.

### Training and validation datasets

The composition of the UK Biobank and EyePACS 10K datasets used in this study is shown in Table 1. The UK Biobank was used for training and internal validation. Participants in the UK Biobank were recruited from a UK general population with 90% non-Hispanic White participants and only 5% self-identified as having diabetes “diagnosed by doctor.” The data from the UK Biobank was accessed via a direct request to the UK Biobank (IRB UOA-86299) and was obtained using approved data management and data transfer protocols. 88,318 retinal images from 51,955 unique participants from the UK Biobank with acceptable quality images were used in this study. This dataset comprises information from individuals above 40 years of age.

**Table 1:**
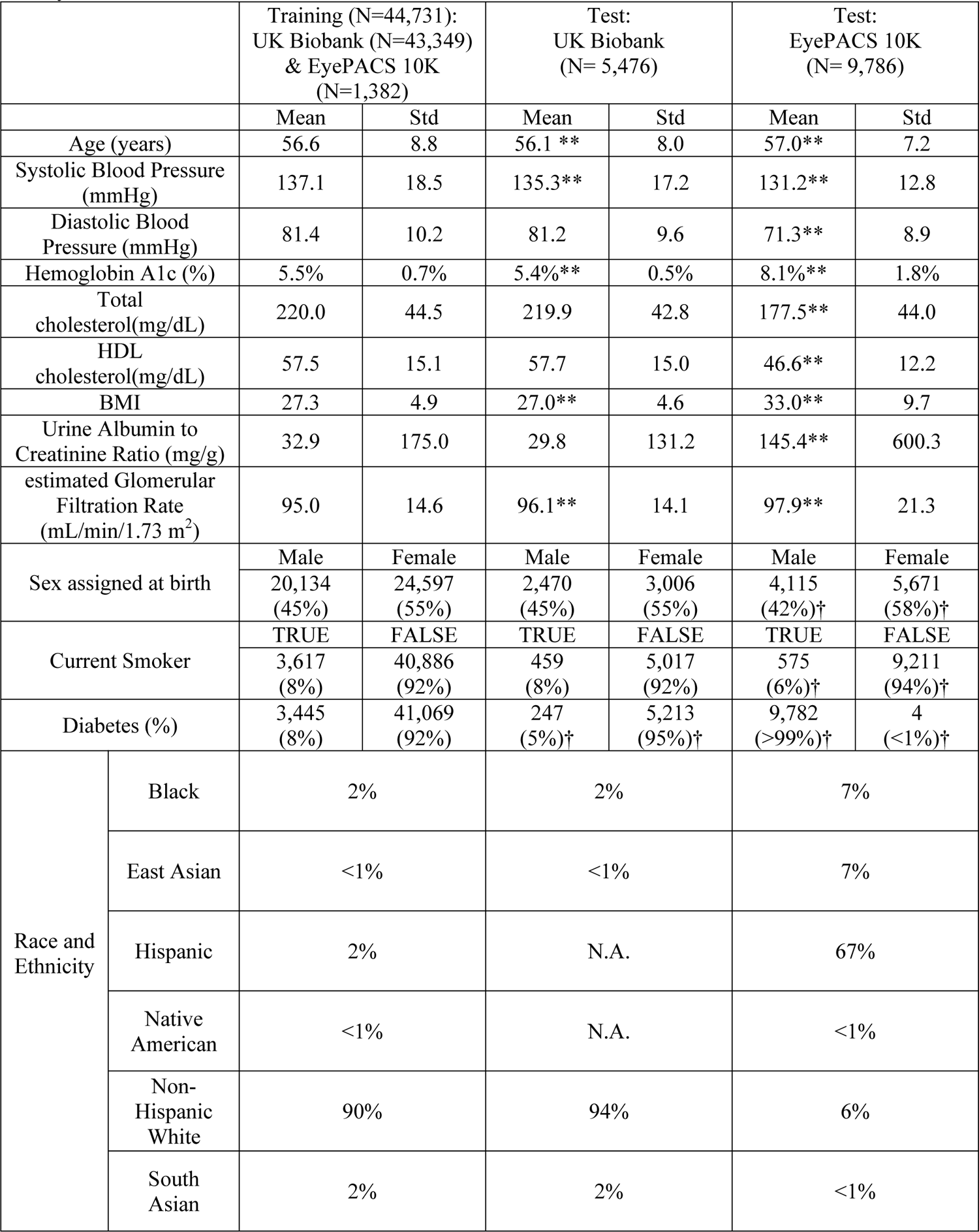
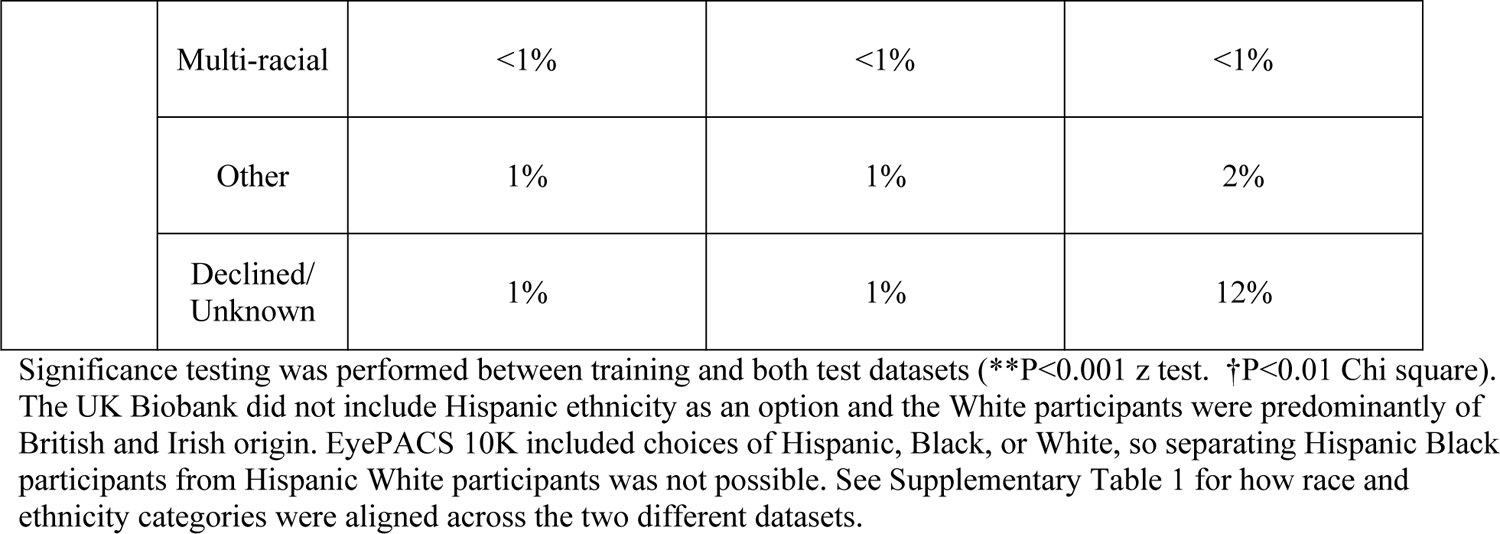
The demographic and risk factor makeup of the training and test datasets used in this study.

|  |  | Training (N=44,731):<br>UK Biobank (N=43,349)<br>& EyePACS 10K<br>(N=1,382) |  | Test:<br>UK Biobank<br>(N= 5,476) |  | Test:<br>EyePACS 10K<br>(N= 9,786) |  |
| --- | --- | --- | --- | --- | --- | --- | --- |
|  |  | Mean | Std | Mean | Std | Mean | Std |
| Age (years) |  | 56.6 | 8.8 | 56.1 ** | 8.0 | 57.0** | 7.2 |
| Systolic Blood Pressure (mmHg) |  | 137.1 | 18.5 | 135.3** | 17.2 | 131.2** | 12.8 |
| Diastolic Blood Pressure (mmHg) |  | 81.4 | 10.2 | 81.2 | 9.6 | 71.3** | 8.9 |
| Hemoglobin A1c (%) |  | 5.5% | 0.7% | 5.4%** | 0.5% | 8.1%** | 1.8% |
| Total cholesterol(mg/dL) |  | 220.0 | 44.5 | 219.9 | 42.8 | 177.5** | 44.0 |
| HDL cholesterol(mg/dL) |  | 57.5 | 15.1 | 57.7 | 15.0 | 46.6** | 12.2 |
| BMI |  | 27.3 | 4.9 | 27.0** | 4.6 | 33.0** | 9.7 |
| Urine Albumin to Creatinine Ratio (mg/g) |  | 32.9 | 175.0 | 29.8 | 131.2 | 145.4** | 600.3 |
| estimated Glomerular Filtration Rate (mL/min/1.73 m<sup>2</sup>) |  | 95.0 | 14.6 | 96.1** | 14.1 | 97.9** | 21.3 |
| Sex assigned at birth |  | Male | Female | Male | Female | Male | Female |
|  |  | 20,134 (45%) | 24,597 (55%) | 2,470 (45%) | 3,006 (55%) | 4,115 (42%)† | 5,671 (58%)† |
| Current Smoker |  | TRUE | FALSE | TRUE | FALSE | TRUE | FALSE |
|  |  | 3,617 (8%) | 40,886 (92%) | 459 (8%) | 5,017 (92%) | 575 (6%)† | 9,211 (94%)† |
| Diabetes (%) |  | 3,445 (8%) | 41,069 (92%) | 247 (5%)† | 5,213 (95%)† | 9,782 (>99%)† | 4 (<1%)† |
| Race and Ethnicity | Black | 2% |  | 2% |  | 7% |  |
|  | East Asian | <1% |  | <1% |  | 7% |  |
|  | Hispanic | 2% |  | N.A. |  | 67% |  |
|  | Native American | <1% |  | N.A. |  | <1% |  |
|  | Non-Hispanic White | 90% |  | 94% |  | 6% |  |
|  | South Asian | 2% |  | 2% |  | <1% |  |

|  |  |  |  |  |
| --- | --- | --- | --- | --- |
|  | Multi-racial | <1% | <1% | <1% |
|  | Other | 1% | 1% | 2% |
|  | Declined/<br>Unknown | 1% | 1% | 12% |

Notably, only about 2% were aged 70 or older, so we limited validation testing to ages 41-70. After further quality screening to ensure compatibility with the deployed ASCVD risk prediction model, we retained 75,271 images from 43,349 participants for training purposes, while 10,976 images from 5,476 participants were preserved for testing the model (Figure 1).

**Figure 1:**
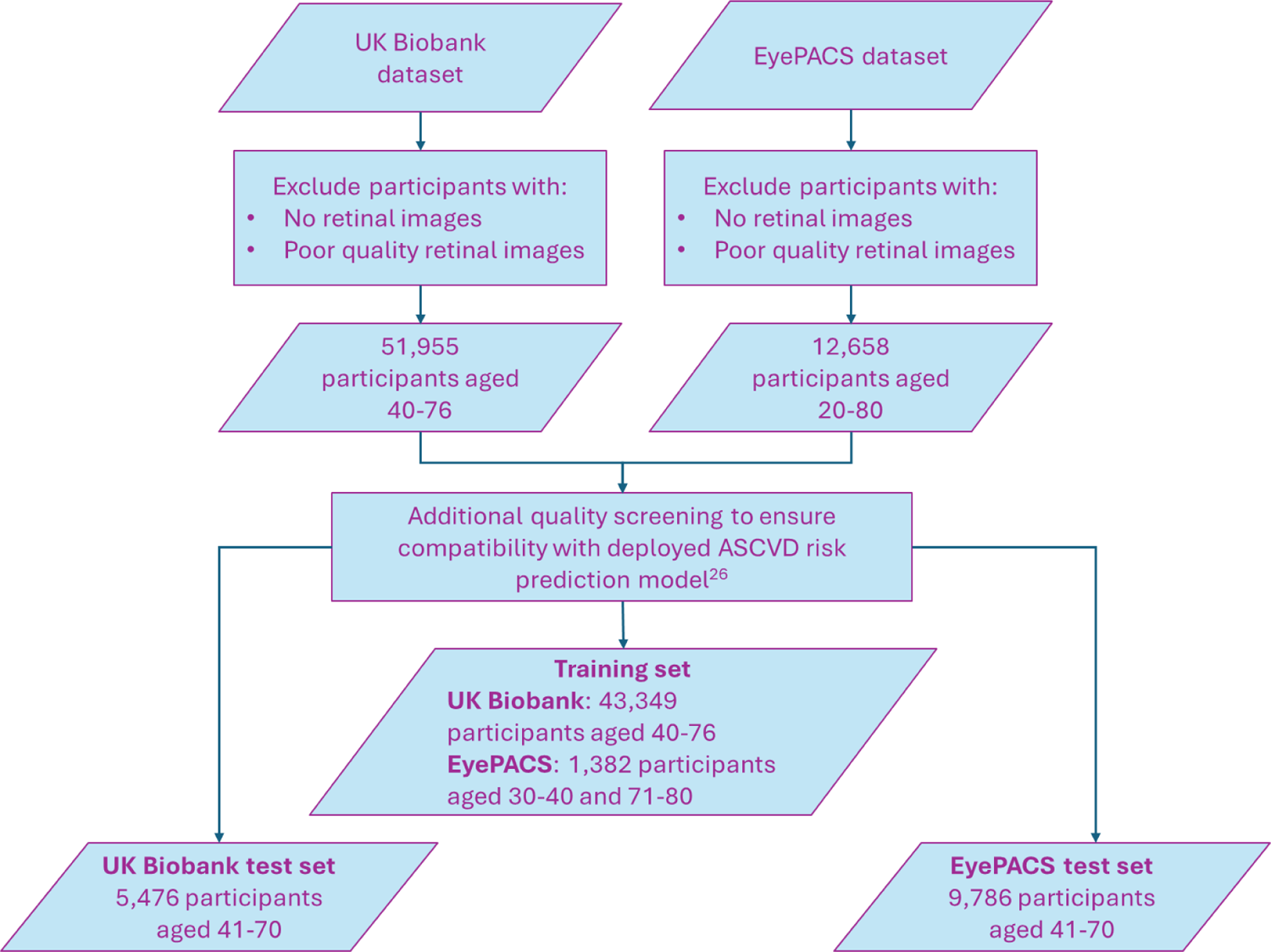
Flow chart illustrating the composition of the training and test datasets used in this study.

The EyePACS 10K dataset (IRB UCB 2017-09-10340) was used for external validation. EyePACS 10K is a diverse, US-based dataset of persons living with diabetes undergoing diabetic retinopathy screening [Table 1]. A subset of 19,856 retinal images from 9,786 individuals in EyePACS 10K was used. These individuals were selected based on having at least one retinal image of good quality from each eye and within the same age range of 41 to 70 years old as tested in UK Biobank.

As the retinal BioAge model requires comparing individuals to peers of higher and lower chronological age, training data for those aged 30-40 and 71-80 were required. UK Biobank is restricted to individuals over 40 years old with only a small representation of those over 70.

Therefore, a limited number of participants from EyePACS 10K who were 40 or younger, or older than 70 were included in the training dataset (Figure 1). Specifically, 853 participants with 1,535 images aged 30-40 (46.3% female, mean age: 36.1 years) and 529 participants with 1,081 images aged 71-80 (65% female, mean age: 73.8 years). Importantly, the separate test datasets for validation analyses in UK Biobank and EyePACS 10K were restricted to individuals aged between 41 and 70 years.

### Retinal BioAge model assessment

To assess the output of the DL model, an individual’s retinal AgeGap (retinal BioAge - chronological age) was calculated. Individuals were grouped in quartiles from the highest to the lowest retinal AgeGap. Then, the prevalence of indicators of CKM syndrome were compared in those in the top vs. bottom quartiles of retinal AgeGap. The following CKM indicators were assessed:

- Blood pressure:

- UK guidelines^30^ define hypertension as SBP≥140mmHg or DBP≥90mmHg, so this threshold was used for analysis of UK Biobank participants.
- US guidelines^31^ define hypertension as SBP≥130mmHg or DBP≥80mmHg, so this threshold was used for analysis of the US-based EyePACS 10K participants.
- Cholesterol:

- Non-HDL cholesterol (Total cholesterol – HDL cholesterol) provides a single lipid measure of risk, with <130 mg/dL considered ideal.^32^
- Kidney function:

- The KDIGO guideline^33^ defines moderate or greater risk chronic kidney disease as estimated GFR <60 mL/min per 1.73m^2^ or urine albumin-to-creatinine ratio (UACR) ≥30 mg/g.
- Diabetes:

- Diabetes is defined as HbA1c ≥6.5% so this threshold was used for analysis of UK Biobank participants.^34^ As the EyePACS 10K participants were known to have diabetes, the threshold for well-controlled diabetes (HbA1c <7.0%) was used.^35^
- Diabetic retinopathy:

- The presence of any diabetic retinopathy (i.e., grade >R0) in either eye was analyzed for EyePACS 10K participants [Supplementary Table 2].^36^

### Data Analysis

Statistical comparison of the prevalence of CKM indicators between the top and bottom retinal AgeGap quartiles was performed by constructing a contingency table followed by the chi-squared test. The prevalence of CKM indicators was also compared using absolute AgeGap values (≥ +2 years vs. ≤ −2 years). Subgroup analysis of male and female participants was also performed. As levels of blood pressure, non-HDL cholesterol, and HbA1c can be impacted by anti-hypertensive, cholesterol-lowering, and diabetes medications, respectively, CKM indicator prevalence was analyzed by medication status (see Supplementary Methods for determination of medication status in UK Biobank and EyePACS 10K).

## Results

Individuals within both the UK Biobank internal validation dataset and the EyePACS 10K external validation dataset were grouped by retinal AgeGap quartiles and the prevalence of CKM indicators assessed [Tables 2, 3]. In UK Biobank, participants within the top retinal AgeGap quartile had a significantly greater prevalence of hypertension (≥ 140/90 mmHg) compared to those in the bottom quartile (36.3% versus 29.0%, p<0.001) [Table 2]. However, there were no significant differences in the prevalence of elevated cholesterol (non-HDL ≥ 130 mg/dL), chronic kidney disease (eGFR < 60 mL/min per 1.73m^2^ or UACR ≥ 30 mg/g), or diabetes (HbA1c ≥ 6.5%) between the low and high retinal AgeGap quartiles.

**Table 2:** UK Biobank: Prevalence of cardiovascular-kidney-metabolic (CKM) syndrome indicators for top and bottom retinal BioAge quartiles.

| CKM indicator | Top Retinal Age Gap Quartile | Bottom Retinal AgeGap Quartile | p-value |
| --- | --- | --- | --- |
| SBP ≥140 mmHg or DBP ≥90 mmHg | 473/1302=36.3% | 329/1134=29.0% | <0.001 |
| Non-HDL Cholesterol ≥130 mg/dL | 1014/1302=77.9% | 889/1134=78.4% | 0.80 |
| eGFR <60 mg/min per 1.73m<sup>2</sup> or UACR ≥30 mg/g | 62/1301=4.8% | 48/1133=4.2% | 0.60 |
| HbA1c≥6.5% | 35/1145=3.1% | 22/1018=2.2% | 0.24 |

SBP – systolic blood pressure, DBP – diastolic blood pressure, HDL – high-density lipoprotein, eGFR – estimated glomerular filtration rate, UACR – urine albumin-creatinine ratio, HbA1c - Hemoglobin A1c.

**Table 3:** EyePACS 10K: Prevalence of cardiovascular-kidney-metabolic (CKM) syndrome indicators for top and bottom retinal BioAge quartiles.

| CKM indicator | Top Retinal AgeGap Quartile | Bottom Retinal AgeGap Quartile | p-value |
| --- | --- | --- | --- |
| SBP ≥130 mmHg or DBP ≥80 mmHg | 1133/2104=53.8% | 1236/2232=55.4% | 0.33 |
| Non-HDL Cholesterol ≥130 mg/dL | 992/1986=49.9% | 930/2162=43.0% | <0.001 |
| eGFR <60 mg/min per 1.73m<sup>2</sup> or UACR ≥30 mg/g | 785/2137=36.7% | 532/2301=23.1% | <0.001 |
| HbA1c≥7.0% | 1622/2120=76.5% | 1371/2284=60.0% | <0.001 |
| DR grade >0 | 1230/2326=52.9% | 193/2415=8.0% | <0.001 |

SBP – systolic blood pressure, DBP – diastolic blood pressure, HDL – high-density lipoprotein, eGFR – estimated glomerular filtration rate, UACR – urine albumin-creatinine ratio, HbA1c - Hemoglobin A1c, DR – diabetic retinopathy.

In contrast, for EyePACS 10K, individuals within the top vs. bottom retinal AgeGap quartiles showed a higher prevalence of elevated cholesterol (49.9% vs. 43.0%, p < 0.001), chronic kidney disease (36.7% vs. 23.1%, p < 0.001), and suboptimally controlled (HbA1c ≥ 7%) diabetes (76.5% versus 60.0%, p < 0.001) [Table 3]. However, there was no significant difference in the prevalence of hypertension (≥130/80 mmHg). Also in EyePACS 10K, individuals within the top retinal AgeGap quartile had significantly higher prevalence of diabetic retinopathy (grade >R0) compared to those individuals in the bottom retinal AgeGap quartile (52.9% versus 8.0%, p < 0.001) [Table 3].

When both UK Biobank and EyePACS 10K were analyzed by absolute AgeGap thresholds, the findings were very similar [Supplementary Tables 3, 4]. Specifically, UK Biobank participants with AgeGap≥+2 years vs. AgeGap≤-2 years had higher prevalence of hypertension [Supplementary Table 3], while EyePACS 10K participants with AgeGap≥+2 years vs. AgeGap≤-2 years had higher prevalence of elevated cholesterol, chronic kidney disease, suboptimally controlled diabetes, and diabetic retinopathy [Supplementary Table 4].

Separate analyses for female and male participants showed similar findings in UK Biobank, with higher prevalence of hypertension in both women and men [Table 4]. In EyePACS 10K, there were several differences based on sex [Table 5]. Higher prevalence of chronic kidney disease, suboptimally controlled diabetes, and diabetic retinopathy were seen in both men and women, as in the overall EyePACS 10K cohort. For elevated non-HDL cholesterol, this prevalence was only significantly higher in women. Interestingly, hypertension prevalence was significantly lower in women and significantly higher in men, whereas there was no significant difference in the overall EyePACS 10K cohort.

**Table 4:** UK Biobank: Prevalence of cardiovascular-kidney-metabolic (CKM) syndrome indicators for top and bottom retinal BioAge quartiles, analyzed by sex at birth.

| CKM Indicator | Sex | Top Retinal AgeGap Quartile | Bottom Retinal AgeGap Quartile | p-value |
| --- | --- | --- | --- | --- |
| SBP ≥140 mmHg or DBP ≥90 mmHg | female | 275/819=33.6% | 174/660=26.4% | 0.003 |
|  | male | 198/483=41.0% | 155/474=32.7% | 0.009 |
| Non-HDL Cholesterol ≥130 mg/dL | female | 620/819=75.7% | 527/660=79.8% | 0.07 |
|  | male | 394/483=81.6% | 362/474=76.4% | 0.06 |
| eGFR <60 mg/min per 1.73m<sup>2</sup> or UACR ≥30 mg/g | female | 39/818=4.8% | 34/659=5.2% | 0.82 |
|  | male | 23/483=4.8% | 14/474=3.0% | 0.20 |
| HbA1c ≥6.5% | female | 15/712=2.1% | 7/591=1.2% | 0.28 |
|  | male | 20/433=4.6% | 15/427=3.5% | 0.52 |

SBP – systolic blood pressure, DBP – diastolic blood pressure, HDL – high-density lipoprotein, eGFR – estimated glomerular filtration rate, UACR – urine albumin-creatinine ratio, HbA1c - Hemoglobin A1c.

**Table 5:** EyePACS 10K: Prevalence of cardiovascular-kidney-metabolic (CKM) syndrome indicators for top and bottom retinal BioAge quartiles, analyzed by sex at birth.

| CKM Indicator | Sex | Top Retinal AgeGap Quartile | Bottom Retinal AgeGap Quartile | p-value |
| --- | --- | --- | --- | --- |
| SBP ≥130 mmHg or DBP ≥80 mmHg | female | 695/1397=49.7% | 654/1140=57.4% | <0.001 |
|  | male | 438/707=62.0% | 582/1092=53.3% | <0.001 |
| Non-HDL Cholesterol ≥130 mg/dL | female | 705/1297=54.4% | 479/1099=43.6% | <0.001 |
|  | male | 287/689=41.7% | 451/1063=42.4% | 0.79 |
| eGFR <60 mg/min per 1.73m<sup>2</sup> or UACR ≥30 mg/g | female | 404/1403=28.8% | 279/1168=23.9% | 0.006 |
|  | male | 381/734=51.9% | 253/1133=22.3% | <0.001 |
| HbA1c ≥7.0% | female | 1038/1395=74.4% | 660/1162=56.8% | <0.001 |
|  | male | 584/725=80.6% | 711/1122=63.4% | <0.001 |
| DR grade >0 | female | 538/1552=34.7% | 87/1229=7.1% | <0.001 |
|  | male | 692/774=89.4% | 106/1186=8.9% | <0.001 |

SBP – systolic blood pressure, DBP – diastolic blood pressure, HDL – high-density lipoprotein, eGFR – estimated glomerular filtration rate, UACR – urine albumin-creatinine ratio, HbA1c - Hemoglobin A1c, DR – diabetic retinopathy.

When medication status was taken into account for the analyses shown in Tables 2 and 3, there were several additional noteworthy findings. In UK Biobank participants, the significantly higher prevalence of hypertension by AgeGap was in the large majority not on blood-pressure-lowering medications [Table 6]. In EyePACS 10K participants, the significantly higher prevalence of elevated cholesterol by AgeGap was in both those on and off lipid-lowering medications. Similarly, the significantly higher prevalence of suboptimally controlled diabetes by AgeGap in EyePACS 10K was in both those on and off anti-hyperglycemic agents [Table 7].

**Table 6:** UK Biobank: Prevalence of cardiovascular-kidney-metabolic (CKM) syndrome indicators for top and bottom retinal BioAge quartiles, based on medication status.

| CKM Indicator | Meds? | Top Retinal AgeGap Quartile | Bottom Retinal AgeGap Quartile | p-value |
| --- | --- | --- | --- | --- |
| SBP ≥140 mmHg<br>or DBP ≥90 mmHg | Y | 36/74=48.6% | 57/120=47.5% | 0.99 |
|  | N | 437/1228=35.6% | 272/1014=26.8% | <0.001 |
| Non-HDL<br>Cholesterol<br>≥130 mg/dL | Y | 40/76=52.6% | 58/129=45.0% | 0.36 |
|  | N | 974/1226=79.4% | 831/1005=82.7% | 0.06 |
| HbA1c≥6.5% | Y | 6/8=75.0% | 2/2=100.0% | 1.0 |
|  | N | 29/1137=2.6% | 20/1016=2.0% | 0.45 |

SBP – systolic blood pressure, DBP – diastolic blood pressure, HDL – high-density lipoprotein, HbA1c - Hemoglobin A1c.

**Table 7:** EyePACS 10K: Prevalence of cardiovascular-kidney-metabolic (CKM) syndrome indicators for top and bottom retinal BioAge quartiles, based on medication status.

| CKM Indicator | Meds? | Top Retinal AgeGap Quartile | Bottom Retinal AgeGap Quartile | p-value |
| --- | --- | --- | --- | --- |
| SBP ≥130 mmHg<br>or DBP ≥80<br>mmHg | Y | 630/986=63.9% | 736/1168=63.0% | 0.71 |
|  | N | 503/1118=45.0% | 500/1064=47.0% | 0.37 |
| Non-HDL<br>Cholesterol<br>≥130 mg/dL | Y | 541/1131=47.8% | 545/1306=41.7% | 0.003 |
|  | N | 451/855=52.7% | 385/856=45.0% | 0.002 |
| HbA1c≥7.0% | Y | 1352/1683=80.3% | 1100/1659=66.3% | <0.001 |
|  | N | 270/437=61.8% | 271/625=43.4% | <0.001 |

SBP – systolic blood pressure, DBP – diastolic blood pressure, HDL – high-density lipoprotein, HbA1c - Hemoglobin A1c.

## Discussion

Our AI analysis of retinal BioAge compared to chronological age in UK and US cohorts found that individuals with a retinal AgeGap in the top quartile vs. bottom quartile had significantly higher prevalence of multiple indicators of CKM syndrome. Specifically for hypertension in the UK Biobank cohort and for elevated cholesterol, chronic kidney disease, suboptimally controlled diabetes, and diabetic retinopathy in the EyePACS 10K cohort. Secondary analyses by sex at birth and medication use did not substantially alter these findings and the findings were similar when absolute retinal AgeGap thresholds were used instead of quartiles. Thus, retinal BioAge appears promising for identifying individuals at higher risk for important cardiovascular, kidney, and metabolic disease indicators, especially in persons living with diabetes.

The finding of higher prevalence of hypertension in both women and men in the higher retinal AgeGap participants in UK Biobank is important given hypertension is the top modifiable risk factor globally for cardiovascular disease and mortality and is often unrecognized and/or uncontrolled.^1,2,31^ We did not find retinal AgeGap predictive of higher prevalence of hypertension in the overall EyePACS 10K cohort, but a much higher % of EyePACS 10K individuals were on anti-hypertensive medications. Also, diabetes is recognized as an important co-morbidity for initiating anti-hypertensive therapy and achieving blood pressure control.^31^ Interestingly, men in the top quartile in EyePACS 10K did have a significantly higher prevalence of hypertension, but women had the opposite finding.

For elevated cholesterol, we focused on non-HDL cholesterol as the best single risk marker derived from a standard lipid profile (apolipoprotein B has been proposed as superior to non-HDL cholesterol but is not routinely available).^37^ Retinal AgeGap did not predict higher prevalence of elevated cholesterol in UK Biobank, where the overall prevalence was high and use of lipid-lowering medication was low. In contrast, retinal AgeGap was predictive of elevated cholesterol in the overall EyePACS 10K cohort, regardless of lipid-lowering medication status. Subanalysis by sex showed this higher prevalence was significant only in women. Notably, guidelines recommend lipid-lowering therapy in persons living with diabetes,^38^ so while the rates of lipid-lowering therapy and achieving non-HDL cholesterol <130 mg/dL were both >50%, and substantially better in EyePACS 10K vs. UK Biobank, these data show further guideline adherence is needed.

Chronic kidney disease is a key component of the CKM syndrome and is often underrecognized and undertreated, leading to kidney failure as well as contributing to adverse cardiometabolic outcomes.^3,39^ In persons living with diabetes in particular, approximately 40% have chronic kidney disease per the KDIGO risk classification, which is based on having either decreased eGFR or increased UACR.^33,40^ Prior retinal AI studies have focused primarily on predicting chronic kidney disease based on eGFR only.^17–19^ Here we show that retinal BioAge is predictive of higher prevalence of chronic kidney disease using the KDIGO criteria (i.e., eGFR and UACR). In contrast, retinal BioAge was not predictive in UK Biobank, where the overall prevalence of chronic kidney disease was very low (∼5%).

For diabetes status, retinal BioAge was not predictive of HbA1c≥6.5% in UK Biobank, where the overall prevalence was very low (∼3%). In contrast, in EyePACS 10K, retinal AgeGap was predictive of suboptimal diabetes control (HbA1c≥7%) for both women and men. Thus, retinal AgeGap may help identify more persons with diabetes who could benefit from better glycemic control.

One of the components of the suite of DL models within the overall retinal BioAge DL model is a diabetic retinopathy feature detector that has been independently trained to grade diabetic retinopathy.^21^ In the top quartile of retinal AgeGap, >50% had diabetic retinopathy (>R0) while in the bottom quartile <10% had diabetic retinopathy. While this higher prevalence remained significant when analyzed by sex, men in the top retinal AgeGap quartile had a 10-fold higher prevalence compared to the bottom retinal AgeGap quartile. Of note, the top quartile of retinal AgeGap also had significantly higher prevalence of referable (≥R2) diabetic retinopathy (45.8% vs. 3.3%, p<0.001). Modjtahedi et al. have recently demonstrated that not only is diabetic retinopathy associated with future risk of ASCVD events and death, but the magnitude of this risk correlates with the severity of retinopathy.^41^ These authors hypothesize that retinopathy could be a surrogate marker of wider vascular disease and conclude that future calculators that are designed to stratify ASCVD risk in diabetic individuals should incorporate detailed retinopathy input information to model this risk more accurately.

The primary strengths of this study include: 1) demonstrating that accelerated retinal BioAge is associated with higher prevalence of important indicators of CKM syndrome and diabetic retinopathy, particularly in a diverse, higher-risk population of people living with diabetes, 2) testing the retinal BioAge model on two very different datasets, including a US-based dataset, and 3) the DL model is based on only retinal photographs and limited demographic data. With over 100 million eye exams performed annually in the US alone, and the guideline recommendation for annual retinal exams in persons living with diabetes, AI-based point-of-care retinal image analysis may provide an important opportunity to help detect indicators for adverse cardiovascular, kidney, and metabolic health.

Hypertension, chronic kidney disease, and diabetes are each indicators of Stage 2 CKM syndrome and call for lifestyle modification and guideline-directed medical therapy, with additional therapies advocated when multiple indicators are present. Non-HDL cholesterol contributes to cardiovascular risk and is included in the latest proposed 10-year cardiovascular risk calculator, along with measurements of blood pressure, kidney function, and diabetes to personalize risk and guide intensity of preventive therapy.^42^ Notably, in the EyePACS 10K cohort, higher AgeGap was associated with a higher prevalence of suboptimal levels of non-HDL cholesterol and HbA1c, regardless of medication status, highlighting an opportunity to improve preventive care.

Prior work has associated biological aging with a wide range of cardiovascular and other diseases, with a recent series of review articles published in the *Journal of the American College of Cardiology*.^23^ Notably, AI models that estimate biological age from retinal images have shown it can also predict all-cause, cardiovascular, and cancer mortality.^27–28^ We have shown previously that our BioAge model, when tested only in the UK Biobank dataset, correlated with shortened leukocyte telomere length and elevated cardiovascular biomarkers.^26^ In the current study, we evaluated the prevalence of actionable indicators of adverse cardiovascular, kidney, and metabolic health and extend the analysis to a more diverse, higher-risk, US-based population. Notably, recent publications also found retinal biological age as a predictor of kidney failure and diabetic retinopathy.^43,44^

A limitation that is in common with all DL models is the generalizability of the current retinal BioAge model as it was trained primarily on the UK Biobank dataset with limited diversity and low rates of chronic kidney disease and diabetes. Encouragingly, the model performed well on the diverse and higher-disease-burden, US-based EyePACS 10K cohort. The retinal BioAge DL model requires training on cases with chronological ages above and below the age range for analysis, so a limited number of EyePACS 10K cases with ages ≤40 and >70 years were used for training only (with >95% of the training dataset from UK Biobank), with the subsequent testing restricted to age 41-70 years. DL models optimally should be validated and replicated on additional external datasets representative of the target population for clinical use. Notably, the information about medication use was quite different between the two datasets, with UK Biobank relying of self-report and with insulin listed as the only diabetes medication.

In conclusion, these results suggest that a retinal BioAge DL algorithm can detect individuals at higher risk for key indicators of CKM syndrome compared to their chronological peer group. As retinal photographs can be captured rapidly and noninvasively, these algorithms could be widely deployed in eye care and primary care settings without significant additional investment.

Moreover, the ease of use makes these technologies particularly relevant to lower-resource settings and thus facilitates the broader detection of high-risk populations. If these results can be replicated, DL models like ours may offer the potential to significantly increase detection and awareness of CKM syndrome indicators and in doing so may ultimately help improve patient outcomes.

## Data Availability

The datasets from UK Biobank and EyePACS are available through those organizations.

## Acknowledgements

None

## References

1. Global Cardiovascular Risk Consortium, Magnussen C, Ojeda FM, Leong DP, Alegre-Diaz J, Amouyel P, Aviles-Santa L, De Bacquer D, Ballantyne CM, Bernabé-Ortiz A, et al. Global Effect of Modifiable Risk Factors on Cardiovascular Disease and Mortality. N. Engl. J. Med. 2023;389:1273–1285.

2. Martin SS, Aday AW, Almarzooq ZI, Anderson CAM, Arora P, Avery CL, Baker-Smith CM, Barone Gibbs B, Beaton AZ, Boehme AK, et al. 2024 Heart Disease and Stroke Statistics: A Report of US and Global Data From the American Heart Association. Circulation. 2024;149:e347–e913.

3. Kidney disease statistics for the United States [Internet]. National Institute of Diabetes and Digestive and Kidney Diseases. 2024 [cited 2024 Jul 10];Available from: https://www.niddk.nih.gov/health-information/health-statistics/kidney-disease

4. Balooch Hasankhani M, Mirzaei H, Karamoozian A. Global trend analysis of diabetes mellitus incidence, mortality, and mortality-to-incidence ratio from 1990 to 2019. Sci. Rep. 2023;13:21908.

5. Ndumele CE, Rangaswami J, Chow SL, Neeland IJ, Tuttle KR, Khan SS, Coresh J, Mathew RO, Baker-Smith CM, Carnethon MR, et al. Cardiovascular-Kidney-Metabolic Health: A Presidential Advisory From the American Heart Association. Circulation. 2023;148:1606– 1635.

6. Ndumele CE, Neeland IJ, Tuttle KR, Chow SL, Mathew RO, Khan SS, Coresh J, Baker-Smith CM, Carnethon MR, Després J-P, et al. A Synopsis of the Evidence for the Science and Clinical Management of Cardiovascular-Kidney-Metabolic (CKM) Syndrome: A Scientific Statement From the American Heart Association. Circulation. 2023;148:1636– 1664.

7. Wagner SK, Fu DJ, Faes L, Liu X, Huemer J, Khalid H, Ferraz D, Korot E, Kelly C, Balaskas K, et al. Insights into Systemic Disease through Retinal Imaging-Based Oculomics. Transl. Vis. Sci. Technol. 2020;9:6.

8. Cheung CY, Biousse V, Keane PA, Schiffrin EL, Wong TY. Hypertensive eye disease. Nat Rev Dis Primers. 2022;8:14.

9. Yang Z, Tan T-E, Shao Y, Wong TY, Li X. Classification of diabetic retinopathy: Past, present and future. Front. Endocrinol. 2022;13:1079217.

10. AI and the retina: Finding patterns of systemic disease [Internet]. American Academy of Ophthalmology. 2021 [cited 2024 Jul 10];Available from: https://www.aao.org/eyenet/article/ai-and-retina-finding-patterns-of-systemic-disease

11. Zhou Y, Chia MA, Wagner SK, Ayhan MS, Williamson DJ, Struyven RR, Liu T, Xu M, Lozano MG, Woodward-Court P, et al. A foundation model for generalizable disease detection from retinal images. Nature. 2023;622:156–163.

12. Poplin R, Varadarajan AV, Blumer K, Liu Y, McConnell MV, Corrado GS, Peng L, Webster DR. Prediction of cardiovascular risk factors from retinal fundus photographs via deep learning. Nat Biomed Eng. 2018;2:158–164.

13. Vaghefi E, Squirrell D, Yang S, An S, Xie L, Durbin MK, Hou H, Marshall J, Shreibati J, McConnell MV, et al. Development and validation of a deep-learning model to predict 10-year atherosclerotic cardiovascular disease risk from retinal images using the UK Biobank and EyePACS 10K datasets. Cardiovasc Digit Health J. 2024;5:59–69.

14. Yi JK, Rim TH, Park S, Kim SS, Kim HC, Lee CJ, Kim H, Lee G, Lim JSG, Tan YY, et al. Cardiovascular disease risk assessment using a deep-learning-based retinal biomarker: a comparison with existing risk scores. Eur Heart J Digit Health. 2023;4:236–244.

15. Zekavat SM, Raghu VK, Trinder M, Ye Y, Koyama S, Honigberg MC, Yu Z, Pampana A, Urbut S, Haidermota S, et al. Deep Learning of the Retina Enables Phenome- and Genome-Wide Analyses of the Microvasculature. Circulation. 2022;145:134–150.

16. Mordi IR, Trucco E, Syed MG, MacGillivray T, Nar A, Huang Y, George G, Hogg S, Radha V, Prathiba V, et al. Prediction of Major Adverse Cardiovascular Events From Retinal, Clinical, and Genomic Data in Individuals With Type 2 Diabetes: A Population Cohort Study. Diabetes Care. 2022;45:710–716.

17. An S, Vaghefi E, Yang S, Xie L, Squirrell D. Examination of alternative eGFR definitions on the performance of deep learning models for detection of chronic kidney disease from fundus photographs. PLoS One. 2023;18:e0295073.

18. Joo YS, Rim TH, Koh HB, Yi J, Kim H, Lee G, Kim YA, Kang S-W, Kim SS, Park JT. Non-invasive chronic kidney disease risk stratification tool derived from retina-based deep learning and clinical factors. NPJ Digit. Med. 2023;6:114.

19. Tan Y, Ma Y, Rao S, Sun X. Performance of deep learning for detection of chronic kidney disease from retinal fundus photographs: A systematic review and meta-analysis. Eur. J. Ophthalmol. 2023;11206721231199848.

20. Abràmoff MD, Lavin PT, Birch M, Shah N, Folk JC. Pivotal trial of an autonomous AI-based diagnostic system for detection of diabetic retinopathy in primary care offices. NPJ Digit. Med. 2018;1:39.

21. Vaghefi E, Yang S, Xie L, Hill S, Schmiedel O, Murphy R, Squirrell D. THEIA^TM^ development, and testing of artificial intelligence-based primary triage of diabetic retinopathy screening images in New Zealand. Diabet. Med. 2021;38:e14386.

22. Comfort A. Test-battery to measure ageing-rate in man. Lancet. 1969;2:1411–1414.

23. Fuster Valentin. Chronological vs Biological Aging. J. Am. Coll. Cardiol. 2024;83:1614– 1618.

24. Putin E, Mamoshina P, Aliper A, Korzinkin M, Moskalev A, Kolosov A, Ostrovskiy A, Cantor C, Vijg J, Zhavoronkov A. Deep biomarkers of human aging: Application of deep neural networks to biomarker development. Aging. 2016;8:1021–1033.

25. Oh HS-H, Rutledge J, Nachun D, Pálovics R, Abiose O, Moran-Losada P, Channappa D, Urey DY, Kim K, Sung YJ, et al. Organ aging signatures in the plasma proteome track health and disease. Nature. 2023;624:164–172.

26. Vaghefi E, An S, Corbett R, Squirrell D. Association of retinal image-based, deep learning cardiac BioAge with telomere length and cardiovascular biomarkers. Optom. Vis. Sci. [Internet]. 2024;Available from: 10.1097/OPX.0000000000002158

27. Nusinovici S, Rim TH, Yu M, Lee G, Tham Y-C, Cheung N, Chong CCY, Da Soh Z, Thakur S, Lee CJ, et al. Retinal photograph-based deep learning predicts biological age, and stratifies morbidity and mortality risk. Age Ageing. 2022;51:afac065.

28. Zhu Z, Shi D, Guankai P, Tan Z, Shang X, Hu W, Liao H, Zhang X, Huang Y, Yu H, et al. Retinal age gap as a predictive biomarker for mortality risk. Br. J. Ophthalmol. 2023;107:547–554.

29. Ahadi S, Wilson KA, Babenko B, McLean CY, Bryant D, Pritchard O, Kumar A, Carrera EM, Lamy R, Stewart JM, et al. Longitudinal fundus imaging and its genome-wide association analysis provide evidence for a human retinal aging clock. Elife. 2023;12:e82364.

30. Boffa RJ, Constanti M, Floyd CN, Wierzbicki AS, Guideline Committee. Hypertension in adults: summary of updated NICE guidance. BMJ. 2019;367:l5310.

31. Whelton Paul K., Carey Robert M., Aronow Wilbert S., Casey Donald E., Collins Karen J., Dennison Himmelfarb Cheryl, DePalma Sondra M., Gidding Samuel, Jamerson Kenneth A., Jones Daniel W., et al. 2017 ACC/AHA/AAPA/ABC/ACPM/AGS/APhA/ASH/ASPC/NMA/PCNA Guideline for the Prevention, Detection, Evaluation, and Management of High Blood Pressure in Adults. J. Am. Coll. Cardiol. 2018;71:e127–e248.

32. Blood Cholesterol: Diagnosis [Internet]. NHLBI, NIH. [cited 2024 Jul 10];Available from: https://www.nhlbi.nih.gov/health/blood-cholesterol/diagnosis

33. Kidney Disease: Improving Global Outcomes (KDIGO) CKD Work Group. KDIGO 2024 Clinical Practice Guideline for the Evaluation and Management of Chronic Kidney Disease. Kidney Int. 2024;105:S117–S314.

34. American Diabetes Association Professional Practice Committee. 2. Diagnosis and Classification of Diabetes: Standards of Care in Diabetes-2024. Diabetes Care. 2024;47:S20–S42.

35. American Diabetes Association Professional Practice Committee. 6. Glycemic Goals and Hypoglycemia: Standards of Care in Diabetes-2024. Diabetes Care. 2024;47:S111–S125.

36. Wilkinson CP, Ferris FL 3rd, Klein RE, Lee PP, Agardh CD, Davis M, Dills D, Kampik A, Pararajasegaram R, Verdaguer JT, et al. Proposed international clinical diabetic retinopathy and diabetic macular edema disease severity scales. Ophthalmology. 2003;110:1677–1682.

37. Sniderman AD, Navar AM, Thanassoulis G. Apolipoprotein B vs Low-Density Lipoprotein Cholesterol and Non-High-Density Lipoprotein Cholesterol as the Primary Measure of Apolipoprotein B Lipoprotein-Related Risk: The Debate Is Over. JAMA Cardiol. 2022;7:257–258.

38. American Diabetes Association Professional Practice Committee. 10. Cardiovascular Disease and Risk Management: Standards of Care in Diabetes-2024. Diabetes Care. 2024;47:S179–S218.

39. Alfego D, Ennis J, Gillespie B, Lewis MJ, Montgomery E, Ferrè S, Vassalotti JA, Letovsky S. Chronic Kidney Disease Testing Among At-Risk Adults in the U.S. Remains Low: Real-World Evidence From a National Laboratory Database. Diabetes Care. 2021;44:2025–2032.

40. Afkarian M, Sachs MC, Kestenbaum B, Hirsch IB, Tuttle KR, Himmelfarb J, de Boer IH. Kidney disease and increased mortality risk in type 2 diabetes. J. Am. Soc. Nephrol. 2013;24:302–308.

41. Modjtahedi BS, Wu J, Luong TQ, Gandhi NK, Fong DS, Chen W. Severity of Diabetic Retinopathy and the Risk of Future Cerebrovascular Disease, Cardiovascular Disease, and All-Cause Mortality. Ophthalmology. 2021;128:1169–1179.

42. Khan SS, Coresh J, Pencina MJ, Ndumele CE, Rangaswami J, Chow SL, Palaniappan LP, Sperling LS, Virani SS, Ho JE, et al. Novel Prediction Equations for Absolute Risk Assessment of Total Cardiovascular Disease Incorporating Cardiovascular-Kidney-Metabolic Health: A Scientific Statement From the American Heart Association. Circulation. 2023;148:1982–2004.

43. Zhang S, Chen R, Wang Y, Hu W, Kiburg KV, Zhang J, Yang X, Yu H, He M, Wang W, et al. Association of Retinal Age Gap and Risk of Kidney Failure: A UK Biobank Study. Am. J. Kidney Dis. 2023;81:537–544.e1.

44. Tang H, Luo N, Zhang X, Huang J, Yang Q, Lin H, Zhang X. Association between biological aging and diabetic retinopathy. Sci. Rep. 2024;14:10123.

